# Impact of routine asymptomatic screening on COVID-19 incidence in a highly vaccinated university population

**DOI:** 10.1101/2021.10.18.21265057

**Authors:** Rebeca C. Falcão, Michael Otterstatter, May A. Ahmed, Michelle Spencer, Sarafa Iyaniwura, Naveed Z. Janjua, Geoff McKee, Michael A. Irvine

## Abstract

**Background:** With the return of in-person classes, an understanding of COVID-19 transmission in vaccinated university campuses is essential. Given the context of high anticipated vaccination rates and other measures, there are outstanding questions of the potential impact of campus-based asymptomatic screening.

**Methods:** We estimated the expected number of cases and hospitalizations in one semester using rates derived for British Columbia (BC), Canada up to September 15^th^, 2021 and age-standardizing to a University population. To estimate the expected number of secondary cases averted due to routine tests of unvaccinated individuals in a BC post-secondary institution, we used a probabilistic model based on the incidence, vaccination effectiveness, vaccination coverage and *R*_0_. We examined multiple scenarios of vaccine coverage, screening frequency, and pre-vaccination *R*_0_.

**Results:** For one 12 week semester, the expected number of cases is 67 per 50,000 for 80% vaccination coverage and 37 per 50,000 for 95% vaccination coverage. Screening of the unvaccinated population averts an expected 6-16 cases per 50,000 at 80% decreasing to 1-2 averted cases per 50,000 at 95% vaccination coverage for weekly to daily screening. Further scenarios can be explored using a web-based application.

**Interpretation:** Routine screening of unvaccinated individuals may be of limited benefit if vaccination coverage is 80% or greater within a university setting.

## Introduction

By April 2020, nearly all post-secondary institutions in Canada had closed their campuses and moved to online classes to reduce the spread of COVID-19, which had by that time become a global pandemic [1, 2]. With the arrival and availability of highly effective vaccines, universities and colleges have largely returned to in-person classes for fall 2021. The reopening of campuses reintroduces frequent close contact among young adults, which can be a driver of rapid spread of coronavirus. To offset this risk, universities, colleges, and other schools allowing in-person classes have implemented protocols, such as mandatory masking, mandatory vaccinations, physical distancing, routine screening, and a hybrid online and in-person class regime. Yet, even with some of these protocols in place, previous reopenings have resulted in spikes in COVID-19 incidence. According to The New York Times, over 700,000 cases of COVID-19 in the US have been linked to university and college campuses by May 26, 2021 [3]. More recently, the number of new infections on US post-secondary campuses seems to be slowly decreasing, and this could be due to vaccination.

In order for universities to plan a safe return to campus, several studies have analyzed the potential for mitigation strategies to control COVID-19 transmission in a university setting [4–14]. Most of these studies have focused on masks mandates, contact tracing, routine testing, and physical distancing. Some utilized social mixing data from contact surveys [1, 9, 15, 16], whereas others relied on simulations of daily behavioural contacts among students [17]. Only a few of these studies have considered scenarios that include a highly vaccinated university population [1, 2, 16, 18, 19].

Many post-secondary institutions implemented asymptomatic testing of COVID-19 during the fall term, yet it is unclear what additional benefit this might offer, especially when some institutions are reporting very high vaccination coverage among students and staff. Another proposed measure, which may pose logistical and ethical challenges, is the routine testing of unvaccinated individuals. In this work, we review previous modelling of COVID-19 transmission in a university setting and highlight the main implications of mass testing to control spread within the context of a vaccinated population. We provide a new probabilistic modelling analysis of routine screening of unvaccinated students on a university campus, based on the example of the University of British Columbia.

## Methods

### Case & hospitalization rates

Expected numbers of cases per 50,000 across one semester (12 weeks) were derived for various levels of vaccination coverage. This number was chosen to reflect the population size of a large university campus. The rolling seven days mean daily case and hospitalization incidence by age group for the province (24 and under, 25-34, 35-44, 45-54, 55-60, 61-65, 66-70, 71 and over) were obtained for September 15^th^, 2021 within BC using the BC Centre for Disease Control (BCCDC) laboratory confirmed and probable COVID-19 case line list. We decided on this date because is near the beginning of term. The hospitalization and case incidence are based on provincial data, and case incidence does vary by region, however, is strongly correlated with vaccination coverage. Rates for vaccinated and un-vaccinated individuals were standardized using population demographic data obtained for the University of British Columbia as an example post-secondary institute and separated into undergraduate, graduate, and staff populations [20, 21]. These rates were then adjusted to generate expected numbers for given vaccination coverage.

### Probabilistic model

We constructed a probabilistic model to determine the expected number of secondary cases averted due to randomly screening unvaccinated individuals within a vaccinated population. The number of secondary cases averted is dependent on the probability of detecting a case during their infectious period and the expected number of secondary cases. The expected number of secondary cases is the vaccine-adjusted effective reproduction number and was calculated incorporating the vaccine coverage, transmission-blocking and onward transmission efficacy of the vaccine, and the pre-vaccination basic reproduction number (*R*_0_). The probability of detection is composed of the probability that a randomly selected individual from the population is infected and the probability of detection given that the individual is infected. Assuming a given testing frequency, this probability is further composed of the total probability of detection at a given time point and the probability that the individual is still infectious at that time point across all time points.

The expected secondary cases averted for a population are then multiplied by the population of un-vaccinated to obtain the total expected cases averted across one generation time. This is then multiplied by the semester time-period to obtain the total cases in one semester. Although non-pharmaceutical interventions (NPIs) were not explicitly included within the model framework, their effects can be observed through variation of the *R*_0_ parameter. The model is fully described in the supplementary information.

### Parameters

Table 1 provides the fixed parameters and the range of the variable parameters that we used for the model. For pre-vaccination *R*_0_, we considered a range from 5 to 9, allowing for increased transmission due to the Delta variant, the expected increase in contacts in the university population, and the potential impacts of NPIs [22–24]. For vaccination coverage, we consider that 80% to 100% of the population may be fully vaccinated, given the high vaccination rates in BC, and the self-reported vaccination rate at the University of British Columbia (UBC). As of September 28, a total of 73,779 members of UBC have completed their vaccination status declaration, where 94.78% of those have declared they are fully vaccinated [25]. The vaccination effectiveness consists of the transmission-blocking efficacy *v*_*e*_ and the reduction in onward transmission *v*_*o*_. We fixed these parameters to 80% and 40%, respectively [26, 27]. The frequency of testing considered for routine screening of unvaccinated individuals was 1 to 14 days. Lastly, we converted the reported serial interval, with a median of 4 days and ranging from 2 to 10 days [28], into a Gamma distribution Γ(*k, θ*) with shape *k* equal to 3.2 and scale *θ* equal to 0.8 describing the serial interval distribution. Hence, the probability of being infected, but not yet transmitting at time *t* is given by Γ(*t*) = Γ(*k, θ*).

**Table 1:**
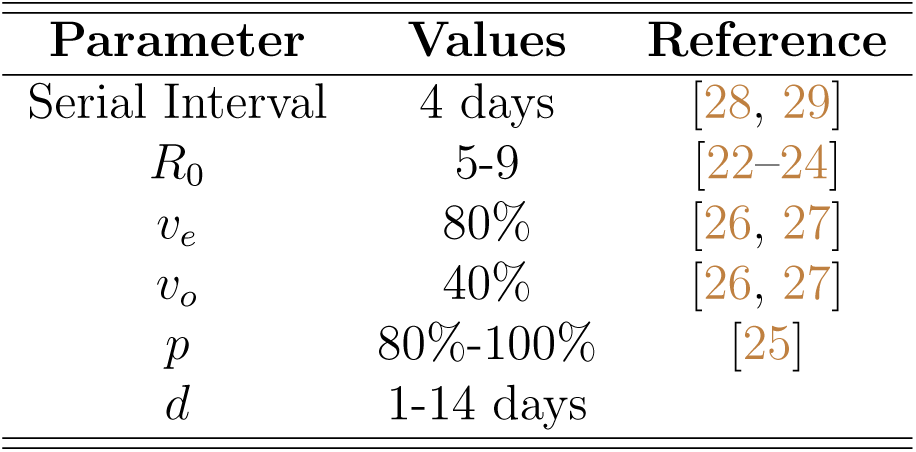
Parameters of probabilistic model

| Parameter | Values | Reference |
| --- | --- | --- |
| Serial Interval | 4 days | [28, 29] |
| $R_0$ | 5-9 | [22–24] |
| $v_e$ | 80% | [26, 27] |
| $v_o$ | 40% | [26, 27] |
| $p$ | 80%-100% | [25] |
| $d$ | 1-14 days | |

The study was reviewed and approved by the University of British Columbia Behavioral Research Ethics Board (No: H20-02097).

## Results

The number of expected cases and hospitalizations during one semester (12 weeks) based on recent epidemiological data is strongly dependent on vaccination coverage. With vaccination coverage of 100%, there are 27 cases per 50,000 expected, compared to 226 cases per 50,000 when coverage is 0%. Expected hospitalizations are 2 per 50,000 in a 100% vaccinated university population compared to 99 per 50,000 in a 0% vaccinated university population (See Table A.1 in supplementary for full details).

Figure 1 displays the expected number of averted secondary cases due to routine screening of unvaccinated students in one 12 week semester per 50000 students, for a testing frequency of 1-14 days (panel 1a: by vaccination coverage, with a pre-vaccination *R*_0_ = 7; panel 1b: by pre-vaccination *R*_0_, with a vaccination coverage of 90%).

**Figure 1:**
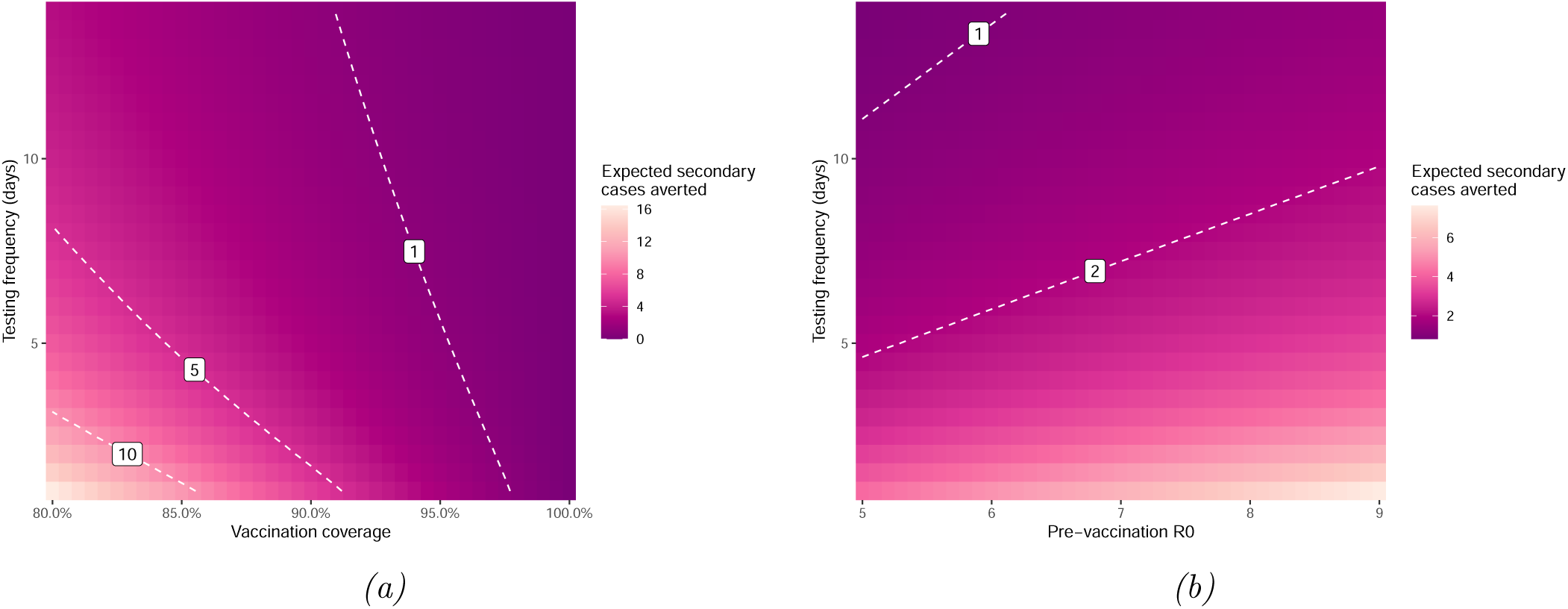
Expected number of secondary cases averted in one 12 week semester per 50000 students. Digits inside box are the number of expected secondary cases averted with parameters values along the curve.**(a)** Change in cases averted by testing frequency and vaccination coverage when pre-vaccination R_0_ = 7. **(b)** Change in cases averted by testing frequency and pre-vaccination R_0_ when vaccination coverage is 90%.

The expected number of secondary cases averted increases with increasing testing frequency and larger pre-vaccination *R*_0_. However, as vaccination coverage increases there are fewer secondary cases that may be prevented by screening. At 80% vaccination coverage and a pre-vaccination *R*_0_ = 7, and given the overall community incidence rate, daily testing averts only an expected 18 secondary cases within a 12 week semester.

Table 2 provides a summary of recent studies, which show that masking and social distancing largely suffice as protective policies in a partially vaccinated population [1, 18]. Zhao et al. conclude that a full university population can safely return to campus if at least 95% are vaccinated [18]. Yang et al. find that a vaccine coverage of > 64% would lead to less than 5% of the university population becoming infected, even with no routine testing [1]. Hambridge et al. find that that frequent testing only confers a substantial reduction in COVID-19 incidence when masking and distancing are not widely adopted, assuming a population with at least 40% vaccine coverage (and a vaccine that is 100% effective) [16]. Both Motta et al. and Junge et al. conclude that mass screening does not provide a substantial reduction in the number of cases when vaccination coverage is high, 90% and 80%, respectively [2, 19].

**Table 2:** Summary of studies that modelled the impact of screening and additional NPIs in a partially vaccinated post-secondary educational institute.

| Study | Transmission Parameters | NPI efficacy | Vaccination efficacy | Vaccination coverage | Implications of mass testing |
| --- | --- | --- | --- | --- | --- |
| Hambridge, 2021 [16] | $R_0$ 1.5,3 and 4.5 | Masking: 85%, distancing: 82% | Assumed 100% | 0 - 60% | With 40% vaccine coverage frequent testing may only confer substantial reduction in incidence when masking and distancing are not widely adopted |
| Zhao, 2021 [18] | Transmission per contact 4.5 - 14% | Masking: 44% (21 - 60), distancing (3ft): 51% (7 - 74), distancing (6ft): 80% (59 - 90) | 95% | 0 - 95% | Outbreaks controlled if 95% are vaccinated without mass testing |
| Yang, 2021 [1] | Transmission probability 3% per contact | Combined 50% | 92 - 95.6% | 50 - 70% | Vaccine coverage > 64% leads to less than 5% of population becoming infected with no testing of asymptomatic individuals |
| Junge, 2021 [2] | $R_0$ 3 | Masking: 50% - 75% | 50 - 80% | 50 - 100% | Mass screening of 25% of population does not substantially change the number of cases if vaccine coverage > 80% |
| Motta, 2021 [19] | Transmission probability 2.6% per contact | interaction multiplier 1, 10, 20 (decrease on NPIs) | 50%, 75%, 90% | 100% | Weekly surveillance testing provides a small reduction in the number of cases if vaccine effectiveness is $\geq 90\%$ |

For other scenarios, the model can be explored using the following web-based application: bccdc.shinyapps.io/university_screening_impact/.

## Interpretation

Routine screening of unvaccinated individuals on a post-secondary campus, as a policy to mitigate COVID-19 spread, poses logistical and ethical challenges. To quantify the impact of such a policy, we developed a probabilistic model to calculate the expectation of the number of secondary cases averted due to routine testing of unvaccinated individuals. In addition, we reviewed previous studies quantifying the impact of different NPIs, including mass testing, on the return to campus of a partially vaccinated population. Our results suggest that routine testing of unvaccinated individuals provides limited additional benefit for the mitigation of COVID-19 spread, given the small expected number of secondary cases that could be averted during a weeks semester with impractical testing frequencies [30–32].

To account for the increase in contacts and the highly transmissible Delta variant, we set a high pre-vaccination *R*_0_ in our model. Yet, with vaccination coverage of at least 80% of the university staff and students, only 18 secondary cases are averted in our analysis during a 12-week semester, when screening is done routinely once per day. These results demonstrate the potential limitations of routine testing among unvaccinated individuals as a policy to reduce new cases when vaccine coverage is high and community incidence is relatively low. Our results agree with previous studies that quantify the impact of mass testing as a strategy to mitigate COVID-19 during return to campus when NPIs are in place and there is partial vaccination coverage. These studies [1, 2, 16, 18, 19] find that mass testing provides a small expected reduction in the number of cases for a population with high vaccination coverage. Masking and physical distancing may be efficient strategies to control the spread of COVID-19 in university settings that already have substantial vaccination coverage [1, 2, 16].

### Limitations

Hospitalizations are underreported in the health authority’s laboratory confirmed and probable COVID-19 case line list, resulting in underestimation of expected hospitalizations cases. In a similar way, cases are also under-reported, and given that probability of being infected in our model is based on daily incidence, it should also be underestimated. A possible solution for those would be to include an estimation of an ascertainment fraction, however higher incidence scenarios can also be explored using the accompanying online application.

Our model does not account for the potential homophily of unvaccinated individuals, which can lead to clusters of cases among the unvaccinated population. Clusters of cases of unvaccinated individuals can potentially be prevented when routine testing unvaccinated individuals, and rapidly isolating the COVID-19 positive cases. Moreover, over-dispersed clusters of cases, a known property of COVID-19 [33–36], is not considered in our model. Over-dispersion would likely increase subsequent secondary cases and the number potentially averted through screening of unvaccinated individuals. However, previous work [1, 2] have found that lower levels of vaccine coverage or efficacy result in right-tail events with a comparatively high number of total cases, for example, values being 5-fold the median (see Figure 1 of Junge et al. [2]). However, they also found that for large vaccination coverage, right-tail events are less likely and with a lower number of cases.

As future work, developing a detailed simulation of COVID-19 dynamics within a university setting could help to more precisely quantify the individual and combined benefits of vaccination and various NPIs, including routine testing. Social contact structure, including potential homophily of unvaccinated individuals and over-dispersion of cases, would be useful additions and could provide a more robust estimation of the number of secondary cases averted.

## Conclusion

For a university campus with high vaccination coverage and low community transmission, the benefits of routine testing of unvaccinated individuals are limited. Other mitigation strategies such as masking and physical distancing may play a more important role in controlling COVID-19 infections compared to mass testing as previous studies have shown [1, 2, 16].

## Data Availability

COVID-19 confirmed case data used in this study is not publicly available. UBC demographics data are available publicly at links given in the manuscript.

## A Appendix

### A.1 Probabilistic model

The expected number of secondary cases averted due to rapid testing an unvaccinated individual in a population with vaccine coverage *p* and pre-vaccination *R*_0_ is given by:

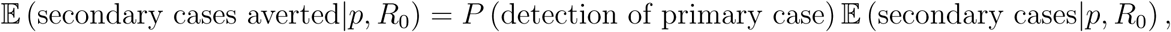

where *P* (detection of primary case) is the probability of testing the primary case, and the expected number of secondary cases that would have arisen due to this detected case is 𝔼 (secondary cases|*p, R*_0_). The expected number of secondary cases in a post-vaccination environment is equal to:

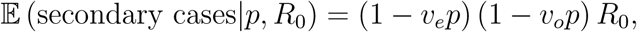

where *v*_*e*_ is the transmission-blocking efficacy of the vaccine, and *v*_*o*_ is the reduction on onward transmission due to vaccine.

The probability of detecting a primary case can be further subdivided into two terms: the probability of being infected, and the probability of being detected given that an individual is infected. Given a testing frequency of *d* days, the probability of detecting a case is modelled by a Poisson process with rate 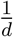. Then, the probability of a primary case not been yet detected by time *t* is given by: 1 − *e*^−*t*/*d*^. The probability of being infected, but not yet transmitting at time *t* is given by Γ(*t*). Thus, we obtain:

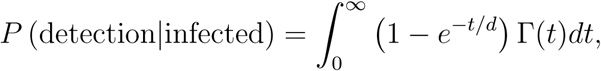

given that the probability of being infected is equal to the incidence on the unvaccinated population, *I*^unv^(*t*), and for a population of size *N* we have *N* (1 − *p*) unvaccinated individuals, the total expected number of secondary cases averted by routine test is given by:

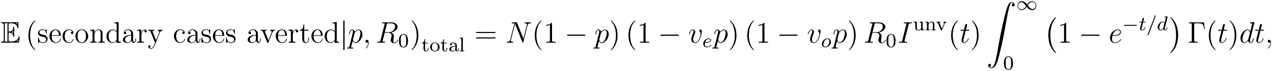

**Table A.1:** Expected number of cases per 50,000 across one semester (12 weeks) by vaccination coverage. Rates were standardized using population demographic data obtained for the University of British Columbia and separated by undergraduate, graduate, and staff populations. The rolling seven day mean daily case incidence by age group were obtained for September 15^th^, 2021 within BC.

| Vaccination coverage | Graduate cases | Staff cases | Undergraduate cases | Total cases | Graduate hospitalizations | Staff hospitalizations | Undergraduate hospitalizations | Total hospitalizations |
| --- | --- | --- | --- | --- | --- | --- | --- | --- |
| 0.0% | 270 | 200 | 225 | 226 | 97 | 255 | 43 | 99 |
| 5.0% | 258 | 191 | 216 | 216 | 93 | 243 | 41 | 95 |
| 10.0% | 246 | 182 | 206 | 206 | 88 | 230 | 39 | 90 |
| 15.0% | 234 | 173 | 196 | 196 | 83 | 218 | 37 | 85 |
| 20.0% | 222 | 164 | 186 | 186 | 78 | 205 | 35 | 80 |
| 25.0% | 209 | 155 | 177 | 176 | 73 | 193 | 33 | 75 |
| 30.0% | 197 | 146 | 167 | 166 | 69 | 180 | 31 | 70 |
| 35.0% | 185 | 137 | 157 | 156 | 64 | 168 | 29 | 65 |
| 40.0% | 173 | 128 | 147 | 146 | 59 | 155 | 27 | 61 |
| 45.0% | 161 | 119 | 137 | 136 | 54 | 142 | 25 | 56 |
| 50.0% | 148 | 110 | 128 | 127 | 49 | 130 | 22 | 51 |
| 55.0% | 136 | 101 | 118 | 117 | 45 | 117 | 20 | 46 |
| 60.0% | 124 | 92 | 108 | 107 | 40 | 105 | 18 | 41 |
| 65.0% | 112 | 83 | 98 | 97 | 35 | 92 | 16 | 36 |
| 70.0% | 99 | 74 | 89 | 87 | 30 | 80 | 14 | 32 |
| 75.0% | 87 | 65 | 79 | 77 | 25 | 67 | 12 | 27 |
| 80.0% | 75 | 56 | 69 | 67 | 21 | 55 | 10 | 22 |
| 85.0% | 63 | 47 | 59 | 57 | 16 | 42 | 8 | 17 |
| 90.0% | 51 | 38 | 50 | 47 | 11 | 30 | 6 | 12 |
| 95.0% | 38 | 29 | 40 | 37 | 6 | 17 | 4 | 7 |
| 100.0% | 26 | 20 | 30 | 27 | 2 | 4 | 2 | 2 |

